# Observation Of Laryngopharyngeal Reflux During Video-Laryngoscopy Was Common Per Our Single Observer Single Institution Experience

**DOI:** 10.1101/2023.06.09.23291219

**Authors:** Ewelina Suchocki, Wael Saasouh, Deepak Gupta

## Abstract

**Background:** Video-laryngoscopy gives an opportunity to objectively document the peri-operative incidence of laryngopharyngeal reflux.

**Materials and Methods:** To quantify the local real-world incidence of laryngopharyngeal reflux, a single observer observed 100 adult patients undergoing elective video-laryngoscopy for their elective surgical procedures in the operating rooms at a single institution.

**Results:** Laryngopharynx in 4% of patients demonstrated visible active refluxing of gastroesophageal contents cranially into laryngopharynx during video-laryngoscopy but none of those underwent suctioning to remove the actively refluxing contents before successfully attempting tracheal intubation.

**Conclusion:** Observed laryngopharyngeal reflux during elective video-laryngoscopy was common among adult patients presenting for elective surgical procedures. The lack of suctioning prior to tracheal intubation warrants consensus discussion and statement regarding whether or not to perform suctioning of visible laryngopharyngeal reflux contents prior to intubation.

## Introduction

The incidence of gastroesophageal (GE) reflux symptoms is rising [1-2] most likely secondary to the rising trends in obesity [3-4]. Consequently, bariatric surgeries are rising as well [5-7] leading to iatrogenic reshaping of GE anatomy. In addition, diabetes and diabetes-related GE symptoms are rising as well [8-9]. Therefore, it has become more important for anesthesiologists to specifically rule out changed GE anatomy and reconfirm perioperative needs per surgical teams. This is especially important before inserting nasogastric or orogastric tubes in awake or anesthetized patients to empty their GE tracts for preventing pulmonary aspiration perioperatively. In this changing environment, video-laryngoscopy has been a life-saver in the operating rooms, on the hospital wards, and outside the hospital in the field [10-12]. Although patients are assumed to be safely devoid of GE reflux if they have adequately followed preoperative fasting guidelines before presenting for their elective surgical procedures, video-laryngoscopy gives an opportunity to objectively document the incidence of laryngopharyngeal reflux that may become the premonitory sign for perioperative pulmonary aspiration secondary to GE reflux contents.

The purpose of this study was to quantify the local real-world incidence [13-14] of laryngopharyngeal reflux observed among our patients undergoing elective video-laryngoscopy.

## Materials and Methods

The institutional review board deemed the present study as non-human participant research. A single observer recorded the following parameters in adult patients undergoing elective video-laryngoscopy for their elective surgical procedures in the operating rooms at a single institution. Only the first electively attempted view with video-laryngoscopy was observed for recording these parameters with the end of observed attempt being either tracheal intubation or returning back to manual/mechanical ventilation. The recorded parameters were:

A. Whether patient was in head-up position
B. Whether manual/mechanical ventilation was performed prior to video-laryngoscopy
C. Whether suctioning was performed prior to video-laryngoscopy
D. Whether larynx was externally manipulated with cricoid pressure or otherwise during video-laryngoscopy
E. Whether laryngopharynx as an opening just posterior to laryngeal cartilages was visible during video-laryngoscopy
  a. If yes, whether anything was actively refluxing cranially from that opening during video-laryngoscopy
    i. If yes, whether suctioning of refluxing material was performed prior to attempting tracheal intubation

## Results

Over a period of six months, co-author ES as the single observer recorded the study parameters among 100 adult patients presenting for elective surgical procedures and undergoing elective video-laryngoscopy. Head-up position during video-laryngoscopy was utilized in all but one patient. Every patient underwent manual ventilation before video-laryngoscopy. Among the 99 patients who underwent video-laryngoscopy in head-up position, the following were the results:

- In 2% patients, video-laryngoscopy failed in its first attempt
- 4% of patients underwent suctioning before video-laryngoscopy
- Laryngopharynx was not visible in 4% of patients during video-laryngoscopy with first attempt at video-laryngoscopy failing among 25% of those patients

Thereafter, among the 95 patients where laryngopharynx was visible during video-laryngoscopy, the following were the results:

- In 1% patients, video-laryngoscopy failed in its first attempt
- 11% of patients had their larynx externally manipulated with cricoid pressure or otherwise during video-laryngoscopy

Finally, among the 85 patients where larynx was not externally manipulated with cricoid pressure or otherwise, the following were the results:

- Laryngopharynx in 4% of patients demonstrated visible active refluxing of GE contents cranially into laryngopharynx during video-laryngoscopy
- None of those three patients underwent suctioning of laryngopharynx to remove the actively refluxing GE contents before successfully attempting tracheal intubation

## Discussion

There were few key findings in this study. Head-up position has become a norm rather than exception among our patients undergoing video-laryngoscopy. However, the degree of head-up position was not recorded in this study which may be highly variable depending on patient’s pre-procedure supine comfort levels as well as provider’s video-laryngoscopy skill comfort levels. This in itself can be a reflection of changing prevalence of premorbid obesity and GE reflux among our patient populations thus making it difficult for our patients to lie flat in bed without any symptoms as well as making it difficult for our providers to perform video-laryngoscopy in supine position without risking intra-procedure hypoxemia or aspiration. As at least 1% incidence denotes common incidence with at least 10% incidence denoting very common incidence [15], failure of first attempts during video-laryngoscopy was still common while external manipulations of larynx with cricoid pressure or otherwise during video-laryngoscopy was very common meaning that second set of hands and skills may still be commonly needed during video-laryngoscopy among our patient populations. Interestingly, laryngopharyngeal reflux was not observed in any patient who underwent external manipulation of larynx with cricoid pressure or otherwise during successful first-attempt intubation with video-laryngoscopy but this was not significantly different (P=0.74; Fisher Exact Test 2×2 contingency table) from common observation of laryngopharyngeal reflux among patients who were successfully intubated without external manipulation of larynx during video-laryngoscopy. Moreover, invisibility of laryngopharynx during video-laryngoscopy risked the failure at first attempt very commonly but that risk was not significantly different (P=0.08; Fisher Exact Test 2×2 contingency table) from common failure of video-laryngoscopy’s first attempt among whom laryngopharynx was visible during video-laryngoscopy. It can be assumed that invisible laryngopharynx may indicate difficult video-laryngoscopy but for that to achieve statistical significance in terms of its implication in failed video-laryngoscopy, a larger study with more patients may be needed. However, our smaller study has quantified that laryngopharyngeal reflux of GE contents was commonly observed during video-laryngoscopy but the actively refluxing contents were never suctioned before successfully attempting tracheal intubation. This may mean that during video-laryngoscopy, the visible laryngeal inlet ready to be intubated may distract providers away from suctioning laryngopharynx prior to or after tracheal intubation.

## Conclusion

Observed laryngopharyngeal reflux during elective video-laryngoscopy was common among adult patients presenting for elective surgical procedures. The lack of suctioning prior to tracheal intubation warrants consensus discussion and statement among providers’ teams and societies regarding whether or not to perform suctioning of visible laryngopharyngeal reflux contents prior to intubation if refluxing contents are not interfering with the visibility of laryngeal inlet.

## Supporting information

Non-Human Participant Reseach Per IRB

## Data Availability

All data produced in the present work are contained in the manuscript

